# Association between lactate to albumin ratio and acute kidney injury in patients with sepsis: a retrospective cohort study from MIMIC-IV database

**DOI:** 10.1101/2024.06.12.24308846

**Authors:** Yaotang Wang, Haixia Yu

## Abstract

**Background:** The mortality rate of sepsis-associated acute kidney injury (S-AKI) is high, yet there is a lack of authoritative prognostic criteria for its outcome. The lactate-to-albumin ratio (LAR) has been associated with mortality in conditions such as sepsis, heart failure, and respiratory failure. However, there is limited research on the relationship between LAR and S-AKI. This study aimed to investigate the association between LAR and the mortality rate of S-AKI.

**Methods:** In our study, we performed a retrospective cohort analysis using the Medical Information Mart for Intensive Care (MIMIC)-IV database. Our primary focus was on determining the occurrence of 30-day, 60-day, and in-hospital mortality as the primary outcomes. To validate the association between LAR and these outcomes, we employed multivariable logistic Cox regression models to calculate hazard ratios.

**Results:** This study included a total of 4793 participants, with a mean age of 63.3 years and a median LAR (log2) of -0.90. After accounting for confounding variables, it was found that patients in the highest LAR (log2) quartile had a higher risk of mortality compared to those in the lowest LAR (log2) quartile. The adjusted hazard ratios for mortality were 1.44 (95% confidence interval [CI]: 1.36∼1.53), 1.38 (95% CI: 1.32∼1.46), and 1.47 (95% CI: 1.39∼1.56) for the highest LAR (log2) quartile, respectively. Moreover, the adjusted hazard ratios for mortality at 30-day, 90-day, and in-hospital intervals were 1.28 (95% CI: 1.23∼1.33), 1.26 (95% CI: 1.21∼1.31), and 1.27 (95% CI: 1.22∼1.32) for each 1 unit increase in LAR, respectively.

**Conclusion:** This study indicates that in septic AKI patients, a higher LAR is associated with an increased risk of all-cause mortality within 30-day, 90-day and in-hospital mortality. This suggests that LAR may serve as an independent risk factor for adverse outcomes in septic AKI patients.

## INTRODUCTION

Sepsis is a global public health challenge, characterized by a dysregulated host response to the infection, leading to a life-threatening organ dysfunction[1, 2]. It affects millions of people worldwide annually, with septic shock and sepsis-related mortality rates of one-third and one-sixth, respectively[3].

Sepsis-associated acute kidney injury (S-AKI) is a major public health disease that is associated with a significant disease burden. S-AKI is an acute dysfunction and organ damage syndrome that may be associated with long-term adverse consequences. Sepsis is the most common cause of acute kidney injury (AKI) in critically ill patients, with sepsis being observed in 40-50% of patients with AKI[4, 5]. Sepsis-associated AKI(S-AKI) is strongly associated with adverse clinical outcomes, as evidenced by a significantly higher mortality rate among septic patients with AKI complications compared to non-AKI patients. This underscores the critical importance of understanding and effectively managing S-AKI in clinical practice[6].In critically ill patients with AKI, S-AKI was associated with a higher risk of in-hospital death and a longer hospital stay compared with AKI from any other cause[7].

Lactic acid is generated through anaerobic glycolysis. It is widely recognized that hyperlactatemia serves as a reliable marker for tissue and organ hypoxia, and heightened levels of lactate have been linked to increased mortality among patients in intensive care units[8].Albumin is synthesized in the liver and is associated with the inflammatory status. With the decrease of serum albumin, mortality in critical ill patients increases substantially[9].

Recent research has shown the predictive significance of the lactate to albumin ratio (LAR) in cases of sepsis and septic shock[10]. Nonetheless, its correlation with 30-day, 90-day, and in-hospital mortality in patients with S-AKI is not yet fully understood.

## MATERIALS AND METHODS

### Data sources

The data utilized for this retrospective analysis was obtained from the freely accessible Medical Information Mart for Intensive Care (MIMIC)-IV-2.2 database, which is open for use by researchers conducting medical research. The authors of this study employed Structured Query Language (SQL) with PostgreSQL to extract pertinent data from the MIMIC IV database. To gain eligibility to access the MIMIC-IV database, the first author completed a training program on the National Institutes of Health (NIH) website, as well as the ‘Data or Specimens Only Research’ and ‘Conflicts of Interest’ examinations (author certification number: 59979404, 59979406, respectively). The Institutional Review Boards of the Massachusetts Institute of Technology exempted the MIMIC-IV database from ethics review due to the anonymization of patient information to safeguard privacy. Our study was conducted in accordance with the principles of the Declaration of Helsinki and adhered to the Strengthening the Reporting of Observational Studies in Epidemiology Reporting Guideline[11].

### Study population

Of the 432,231 admissions in the MIMIC-IV database, 73,181 were ICU admissions. In total, 58201 patients were first hospitalized and first ICU admitted[12].

To screen participants in the study, participants who met the following criteria were included in the study: (1) participants diagnosed with sepsis-3; (2) age 18 years; and (3) first admission to the hospital and intensive care unit (ICU). According to the Sepsis-3 criteria[13], sepsis was defined as a confirmed or suspected infection within 24 hours of ICU admission with a continuous organ failure assessment (SOFA) score of 2.

The following patients were excluded after further screening: (1) patients who did not meet the sepsis-3 definition; (2) only first admission records were kept for repeated admissions; (3) lack of 24-hour blood lactate and serum albumin measurements; and (4) patients who did not progress to AKI seven days after ICU admission within 7 days.

### Date extraction

We used the Navicat Premium software (version 16) to extract the clinical data from the MIMIC-IV. Clinical data included age, sex, length of stay, ICU stay, 30,90 days and in-hospital mortality, heart rate, systolic pressure, respiration, oxygen saturation, 6h urine output, red cell distribution width, hemoglobin, platelets, leukocytes, anion gap, chloride, sodium and potassium ions, INR. Comorbidities included myocardial infraction,renal disease,charlson comorbidity index,SOFA score,AKI stage. Treatment measures included the use of anti-infection, vasopressor, diuretic. In this study, anti-infective treatments include penicillin, cephalosporins, quinolones, premium antibiotics (imipenem, exopenem). The vasopressor included norepinephrine, epinephrine, vasopressin, dopamine, dobutamine, interhydroxylamine, and isoproterenol. Diuretics here represent loop diuretics, thiazide diuretics and potassium-sparing diuretics and the new diuretic toravaptan. Comorbidities were determined according to the 9th and 10th Edition Clinical revision codes.

### Exposure

The exposure factor LAR was equal to lactate divided by albumin. LAR was transformed into LAR(log_2_). We divided LAR(log_2_) into three levels according to quartile distances as follows: tertile 1, LAR(log_2_) < -1.46; tertile 2, -1.46 ≤ LAR(log_2_) < -0.20; tertile 3, LAR(log_2_) ≥ -0.20.

### Study endpoints

In this study, the outcome was 30-day, 90-day and in-hospital mortality.

### Statistical analysis

The participants were categorized into three groups based on LAR(log_2_) quartiles for descriptive analysis. Continuous variables were reported as mean ± SD or median (IQR), and group differences were assessed using ANOVA or the Kruskal-Wallis rank sum test. Categorical variables were presented as frequencies or percentages and analyzed using Chi-square or Fisher test. Before analysis, all LAR data was log_2_ transformed to meet normality assumptions.

Covariates with greater than 10% change in initial regression coefficients were selected based on previous reports and incremental increases or elimination of covariates in the basic and full models. The variance extension factor (VIF) is used to determine whether a collinear relationship exists. A VIF> 2 indicates a collinear relationship. Hazard ratios (HR) and 95% confidence intervals (CI) for the different LAR levels and clinical outcomes were calculated using multivariable cox regression analysis. In Model 1, no adjustment for covariates. Model 2 adjusted for age,sex,heart rate, SBP,respiratory rate, hematocrit, hemoglobin, platelets. Model 3 included model 2 plus AG, chloride, sodium, potassium, INR. Model 4 included model 3 plus myocardial infraction,congestion heart failure, renal disease,SOFA, vasopressor agent. Restricted cubic spline (RCS) regression models were used to analyze continuous LAR and LAR (log2) data to explore the association of LAR with the occurrence of mortality within ICU admission. The spline models included all the same covariates as the multivariable cox regression model 4. Forest plots were generated to illustrate the subgroup analysis and to assess the interaction between LAR and mortality in septic AKI patients by age, sex, SOFA score, AKI stage and different comorbidities classification.

A p value less than 0.001 was considered statistically significant to indicate statistical significance, and all tests were two-sided. All the analyses were performed with the Statistical Software(version 4.2.2, http://www.R-project.org, The R Foundation) and a Free Statistical analysis platform (version 1.9.2, Beijing, China)[14].

## RESULTS

### Baseline characteristics of the study subjects

A total of 73,181 adult patients were admitted to the ICU in the MIMIC-IV v 2.2 database, and 4793 septic patients were enrolled for this study according to the inclusion and exclusion criteria in the method section. The study flow chart is shown in Figure 1. The demographic sample characteristics of all participants are shown in Table 1. The mean age of the patients was 63 years, including 2833 (59.1) males and 1960 (40.9) females. The median baseline value of the LAR (log2) was-0.90 (-1.46, -0.20). By LAR quartiles, participants were divided into three groups: tertile 1, LAR (log2) <-1.46, tertile 2, -1.46≤LAR (log2) <-0.20; tertile 3, LAR (log2)≥-0.20. Patients with higher LAR (log2) had higher heart rate, respiratory rate, anion gap, INR, Charlson comorbidity index and Sofa score, but length of hospital stay, ICU stay, systolic blood pressure, diastolic blood pressure, body temperature, SpO_2_, urine output, erythrocyte distribution width, and lower platelets. In addition, patients with the highest quartile of LAR (log_2_) were more likely to be anti-infective, pressors and diuretics and more likely to have comorbid malignancy (Figure 2).

**Figure 1.**
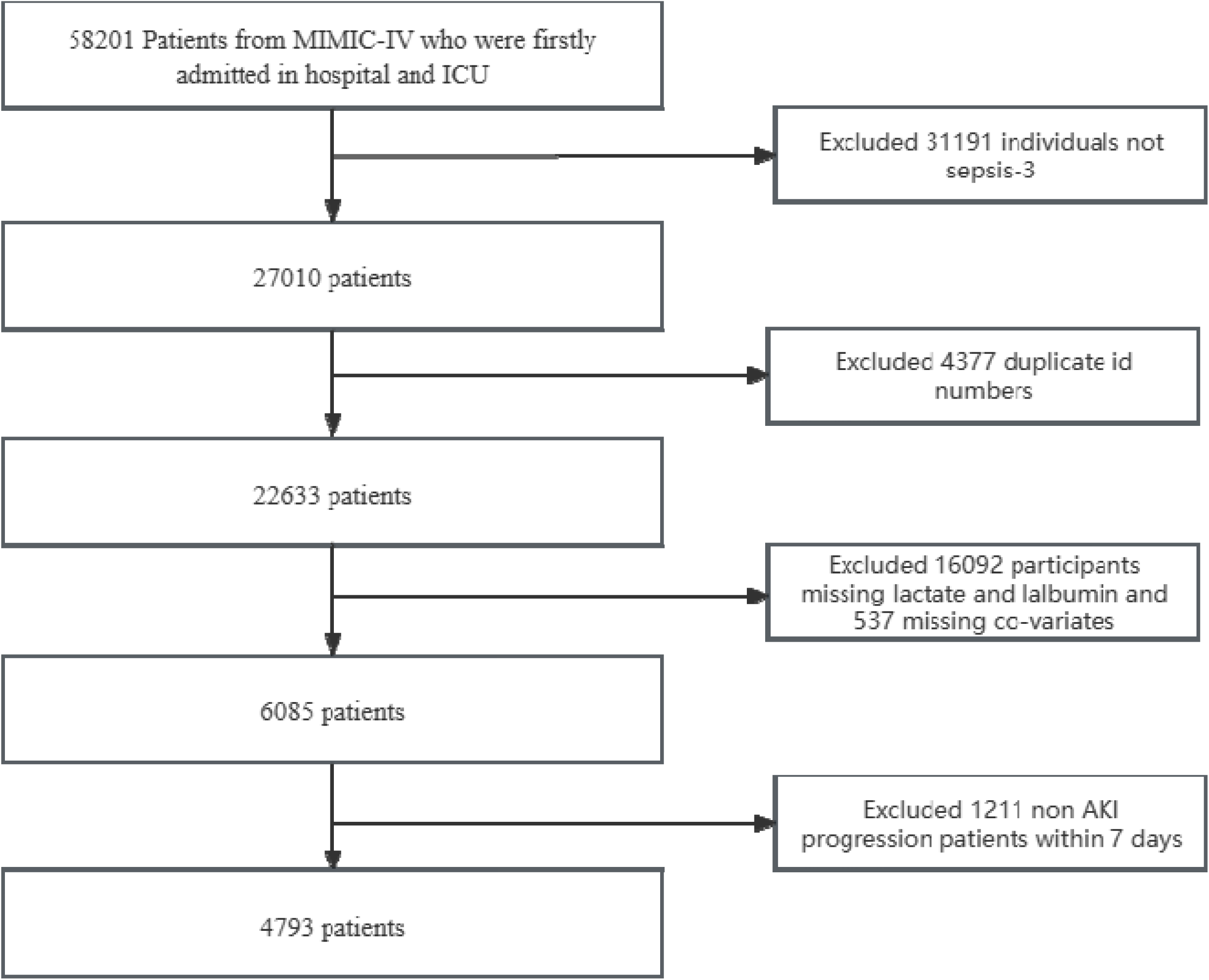
Flow chart

**Table 1.**
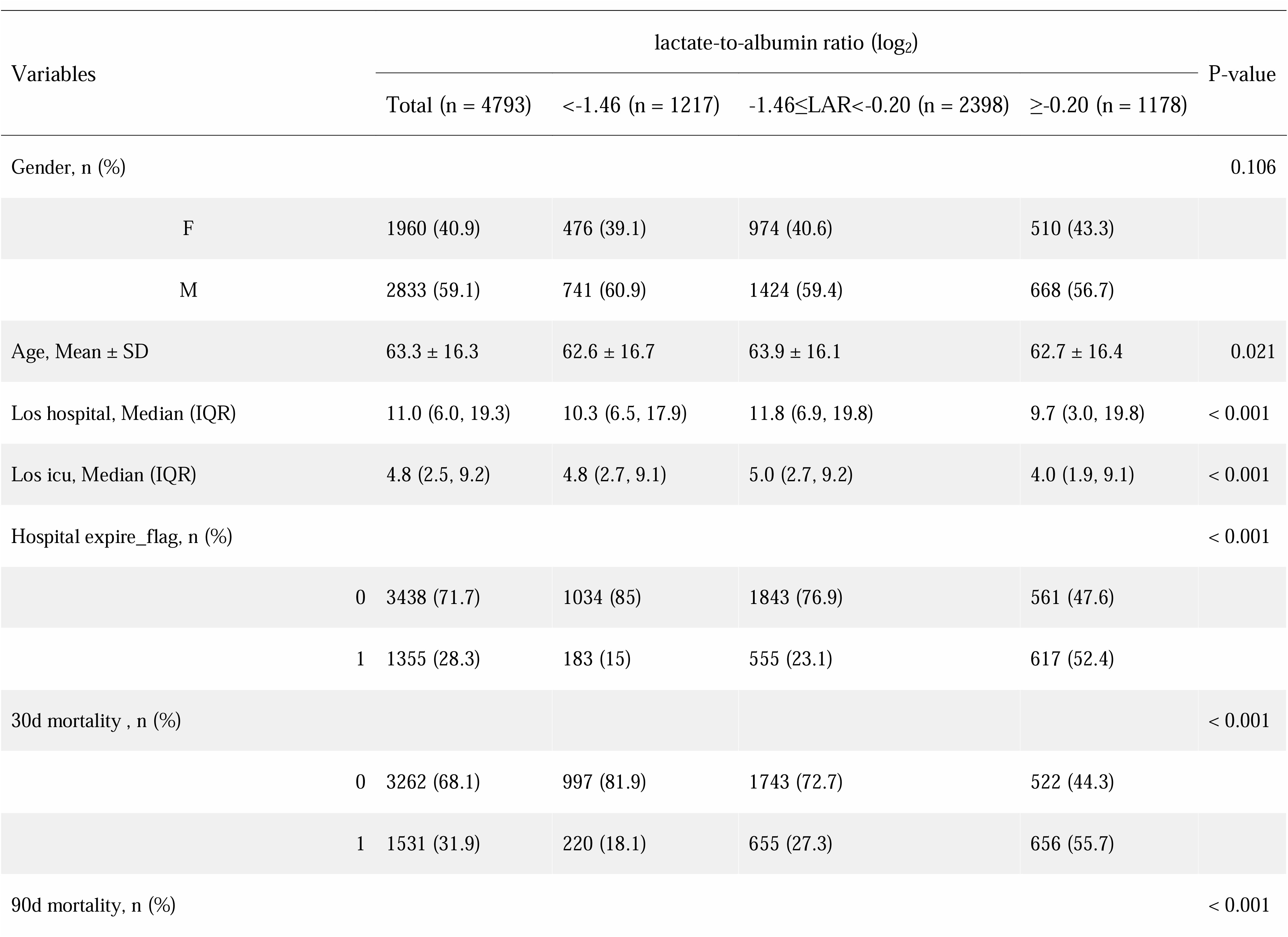
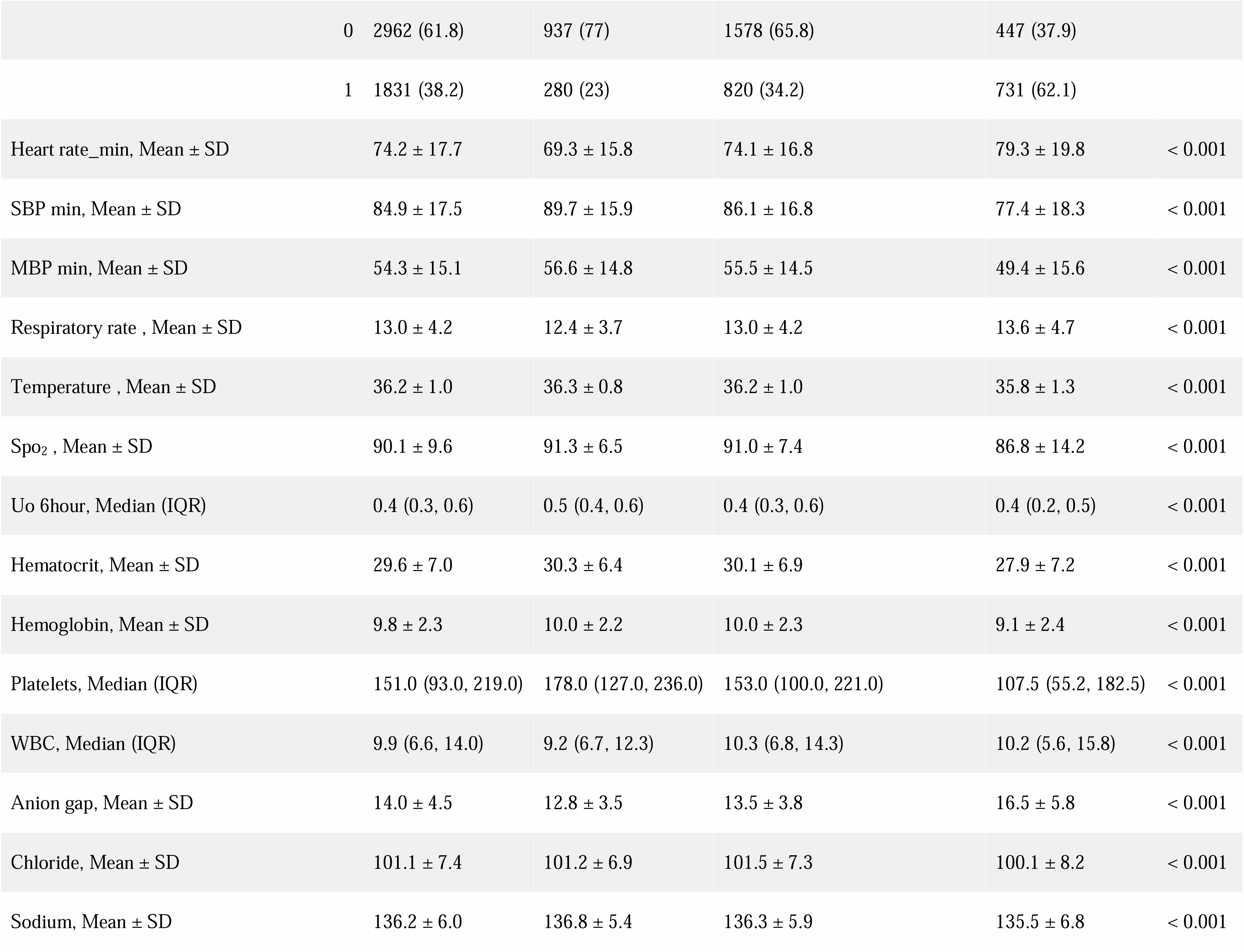
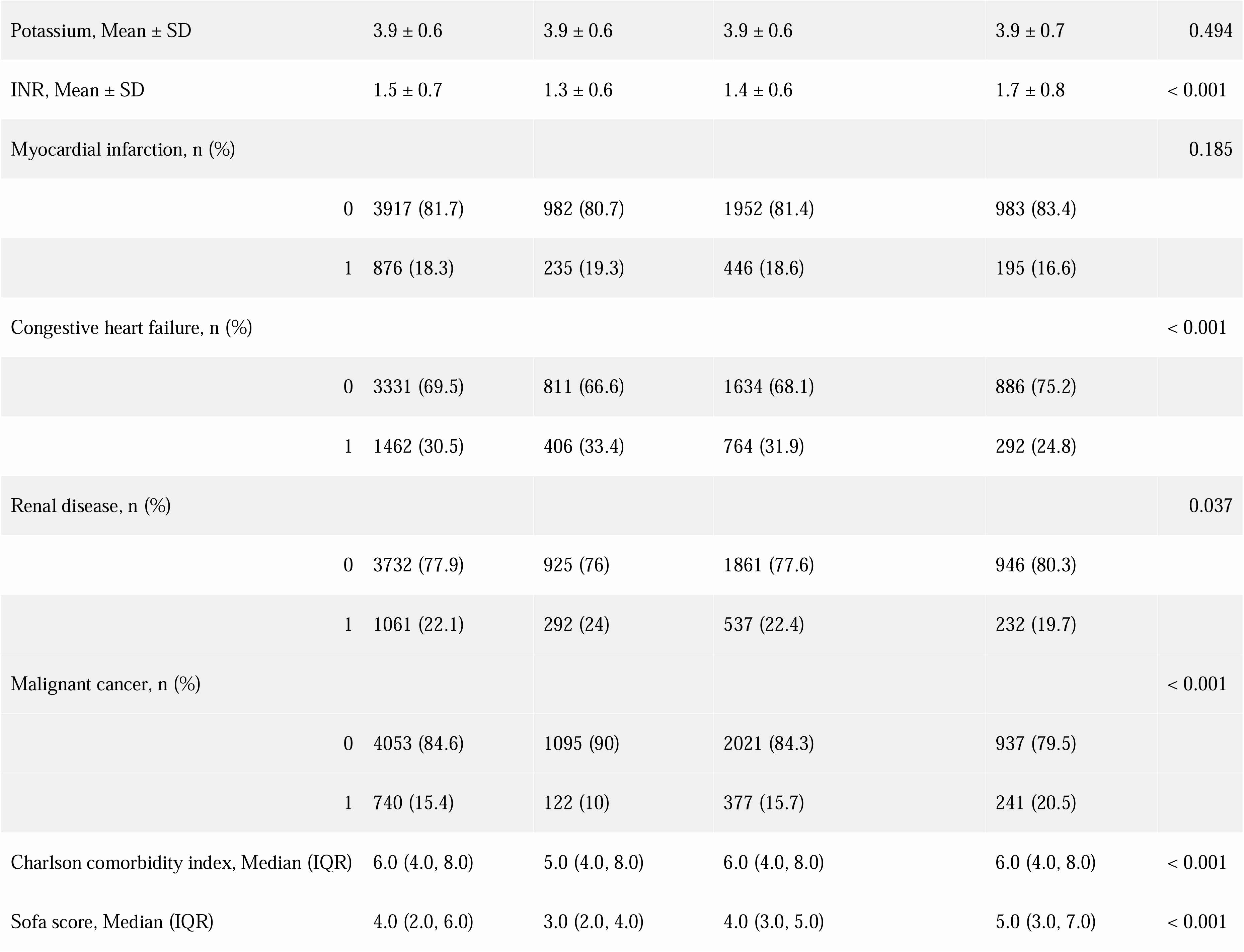
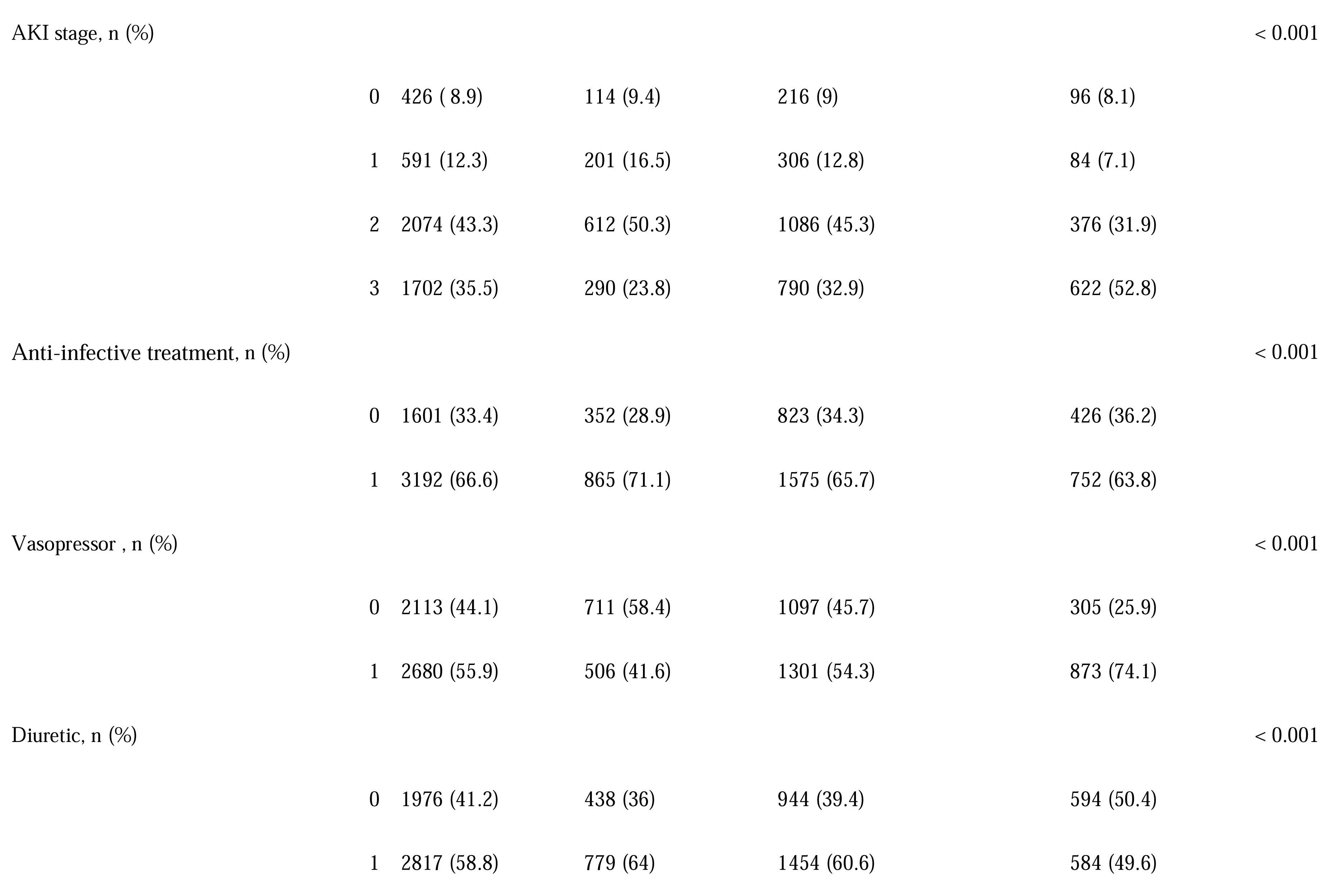
Baseline Characteristics of the enrolled subjects based on LAR tertiles.

**Table 2.** Association of LAR and clinical outcomes in patients with S-AKI.

| Table2 Association of LAR and clinical outcomes in patients with S-AKI |  |  |  |  |  |
| --- | --- | --- | --- | --- | --- |
|  | Variable | Model 1 | Model 2 | Model 3 | Model 4 |

|  | LAR | HR (95% CI) | P-Value | HR (95% CI) | P-Value | HR (95% CI) | P-Value | HR (95% CI) | P-Value |
| --- | --- | --- | --- | --- | --- | --- | --- | --- | --- |
| 30-day mortality | Per 1 unit | 1.44 (1.4~1.47) | <0.001 | 1.39<br>(1.35~1.44) | <0.001 | 1.3 (1.25~1.35) | <0.001 | 1.28<br>(1.23~1.33) | <0.001 |
|  | Per log<sub>2</sub> unit | 1.7 (1.63~1.78) | <0.001 | 1.64<br>(1.56~1.72) | <0.001 | 1.48 (1.4~1.56) | <0.001 | 1.44<br>(1.36~1.53) | <0.001 |
|  | Tertile1 | 1(Ref) |  | 1(Ref) |  | 1(Ref) |  | 1(Ref) |  |
|  | Tertile2 | 1.43<br>(1.23~1.67) | <0.001 | 1.34<br>(1.15~1.56) | <0.001 | 1.29<br>(1.11~1.51) | 0.001 | 1.26 (1.08~1.47) | 0.004 |
|  | Tertile3 | 3.38 (2.9~3.93) | <0.001 | 2.96<br>(2.52~3.48) | <0.001 | 2.4 (2.03~2.83) | <0.001 | 2.24 (1.89~2.66) | <0.001 |
| 90-day mortality | Per 1 unit | 1.38 (1.34~1.42) | <0.001 | 1.34 (1.3~1.38) | <0.001 | 1.27 (1.23~1.32) | <0.001 | 1.26 (1.21~1.31) | <0.001 |
|  | Per log<sub>2</sub> unit | 1.56 (1.5~1.63) | <0.001 | 1.53 (1.46~1.6) | <0.001 | 1.41 (1.34~1.48) | <0.001 | 1.38 (1.32~1.46) | <0.001 |
|  | Tertile1 | 1(Ref) |  | 1(Ref) |  | 1(Ref) |  | 1(Ref) |  |
|  | Tertile2 | 1.39 (1.21~1.59) | <0.001 | 1.32 (1.15~1.52) | <0.001 | 1.28 (1.12~1.47) | <0.001 | 1.26 (1.1~1.45) | 0.001 |
|  | Tertile3 | 2.84 (2.48~3.27) | <0.001 | 2.61 (2.26~3.03) | <0.001 | 2.2 (1.89~2.56) | <0.001 | 2.12 (1.82~2.47) | <0.001 |
| In-hospital<br>mortality | Per 1 unit | 1.44 (1.4~1.48) | <0.001 | 1.39 (1.34~1.43) | <0.001 | 1.29 (1.25~1.34) | <0.001 | 1.27 (1.22~1.32) | <0.001 |
|  | Per log<sub>2</sub> unit | 1.77 (1.69~1.86) | <0.001 | 1.69 (1.6~1.78) | <0.001 | 1.51 (1.43~1.6) | <0.001 | 1.47 (1.39~1.56) | <0.001 |
|  | Tertile1 | 1(Ref) |  | 1(Ref) |  | 1(Ref) |  | 1(Ref) |  |
|  | Tertile2 | 1.45 (1.23~1.72) | <0.001 | 1.35 (1.14~1.6) | 0.001 | 1.29 (1.09~1.53) | 0.003 | 1.25 (1.05~1.48) | 0.011 |
|  | Tertile3 | 3.77 (3.19~4.44) | <0.001 | 3.18 (2.67~3.78) | <0.001 | 2.52 (2.11~3.02) | <0.001 | 2.35 (1.96~2.82) | <0.001 |

Note:
Model 1
No co-variates adjusted
Model 2
Adjusted for age,sex,heart rate, SBP,respiratory rate, hematocrit, hemoglobin, platelets
Model 3
Model 2+(AG, chloride, sodium, potassium, INR)
Model 4
Model 3+(myocardial infraction,congestion heart failure, renal disease,SOFA, vasopressor agent)

### Association between LAR and clinical outcomes

1531,1831 and 1355 patients died at 30 days of ICU admission, 90 days and during hospitalization, respectively. Patients with higher LAR(log_2_) had higher 90-day mortality, ranging from 23% (tertile 1) of patients with LAR(log_2_) <-1.46 to 62.1% (tertile 3) of patients with LAR(log_2_) ≥ -0.20. A similar trend was observed between the 30-day death and LAR (log_2_) incidence.

Multivariate cox regression A significant association between LAR and between LAR (log2) and death in sepsis patients (Table 3). When analyzed as a continuous variable, In the univariate analysis, LAR with 30 days, 90-day death and in-hospital death (HR: 1.44, 95%CI:1.4∼1.47, HR:1.38, 95%CI:1.34∼1.42, HR:1.44, 95%CI: 1.4∼1.48), In Model 2 (HR: 1.39, 95%CI:1.35∼1.44, HR:1.34, 95%CI:1.3∼1.38, HR:1.39, 95%CI1.34∼1.43), Model 3 (HR: 1.3, 95%CI:1.25∼1.35, HR:1.27, 95%CI: 1.23∼1.32, HR:1.29, 95%CI: 1.25∼1.34) and Model 4 (HR: 1.28, 95%CI:1.23∼1.33, HR:1.26, 95%CI:1.21∼1.31, HR: 1.27 95%CI: 1.22∼1.32), After adjustment for the confounding variables. When analyzed by LAR (log2) tertiles, patients with higher LAR tertiles (HR: 3.38,95%CI: 2.9 to 3.93) died higher than the lowest LAR tertile at 30 days). These correlations remained significant in models 2, these 3 and 4 after adjusting for potential covariates. Similar results were observed between the 90-day death and the in-hospital death. After adjusting for confounding variables, the RCS analysis showed that the association between LAR and 30-day, 90-day, and in-hospital death was nonlinear (non-linear P <0.001) (Figure 2a, 2b, 2c). With LAR <0.52, the risk of 30 days, 90 days and hospitalization death increased rapidly with LAR (HR 2.037,95%CI=1.641,2.528; HR: 3.868,95%CI=1.571,9.524; HR 2.583 95%CI=0.848,7.865). Risk increased relatively slowly when the LAR exceeded 0.52 (HR 1.254,95%CI=1.184,1.327; HR: 1.223,95%CI=1.157,1.29).

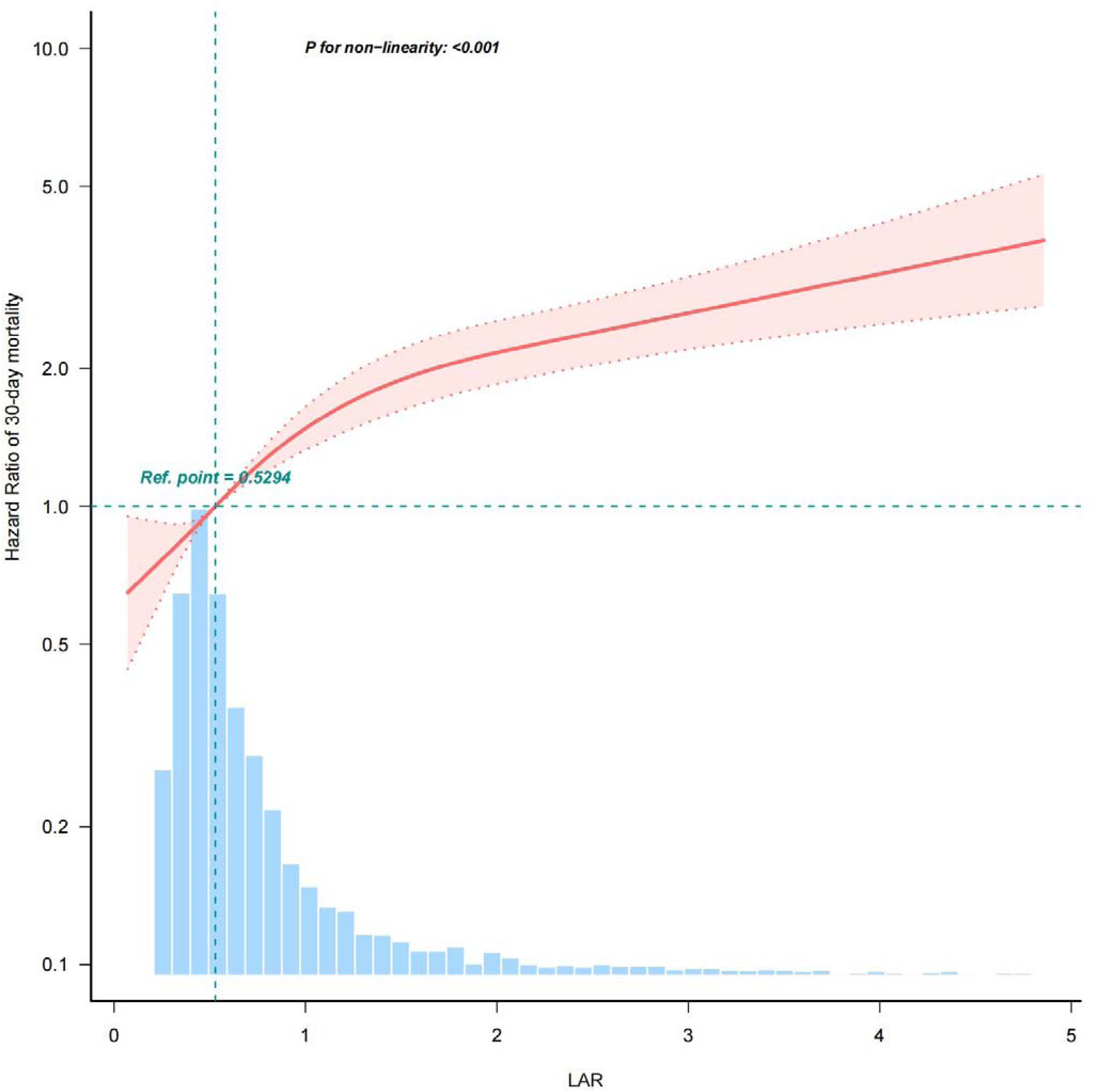
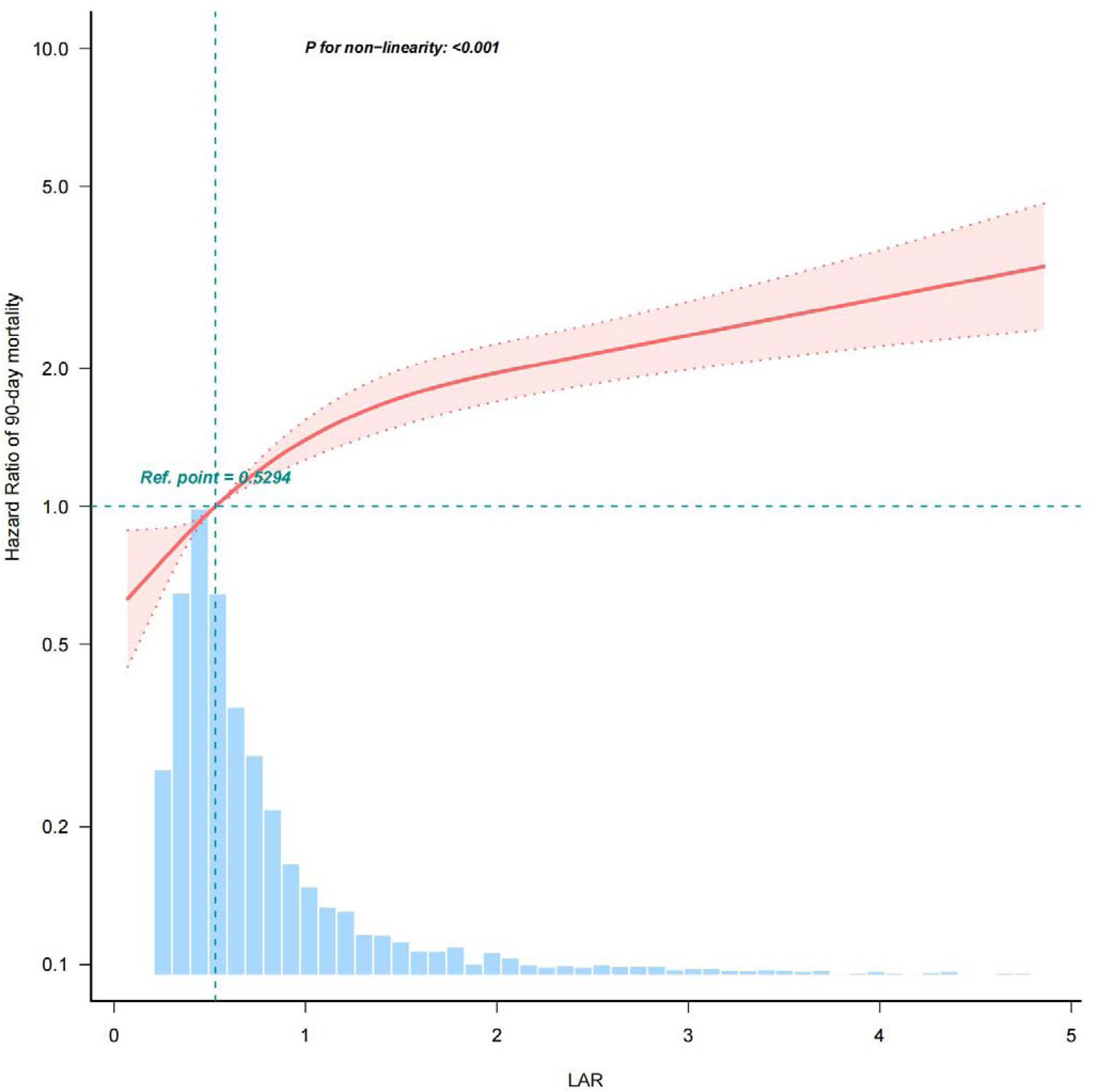
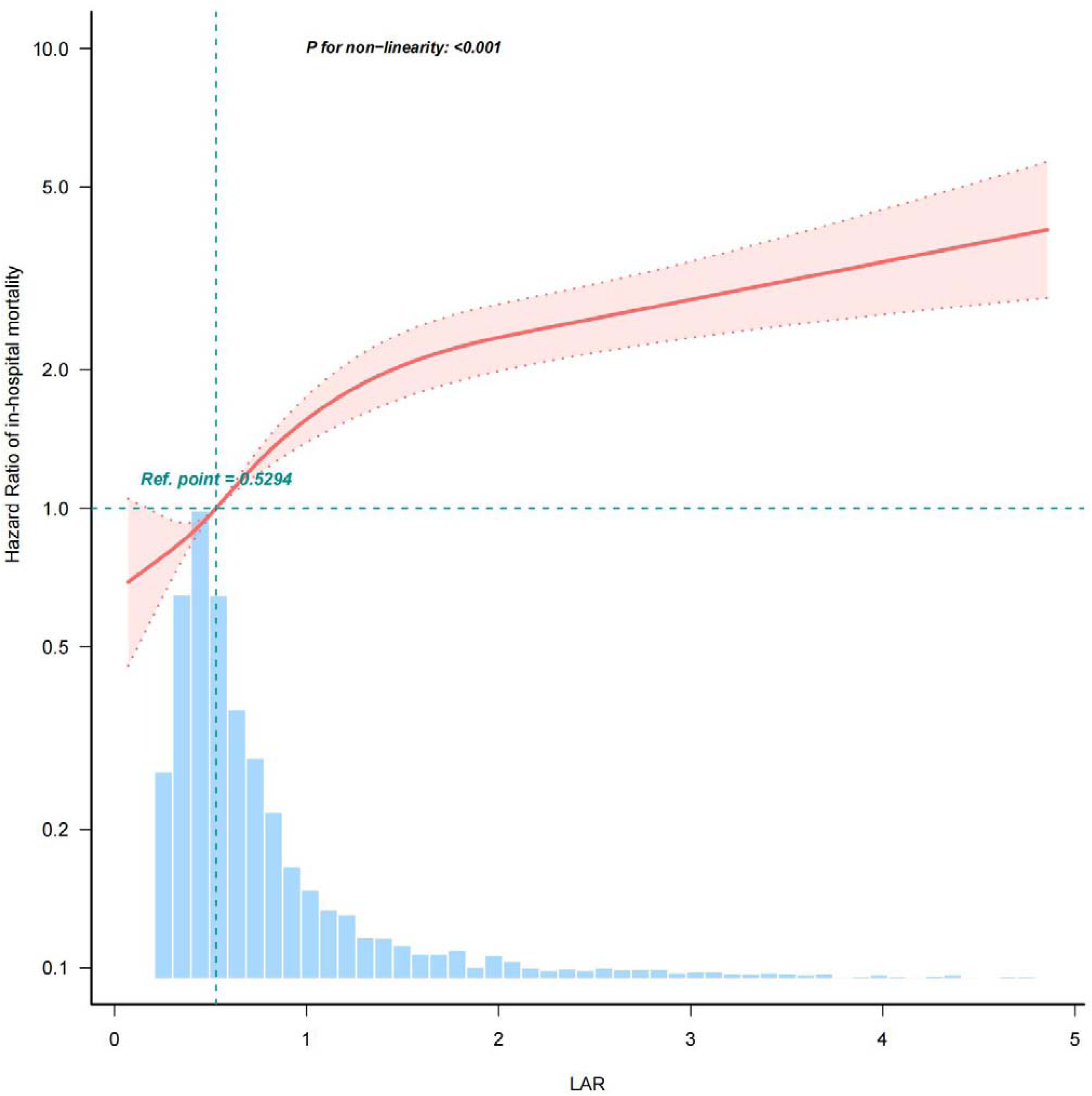
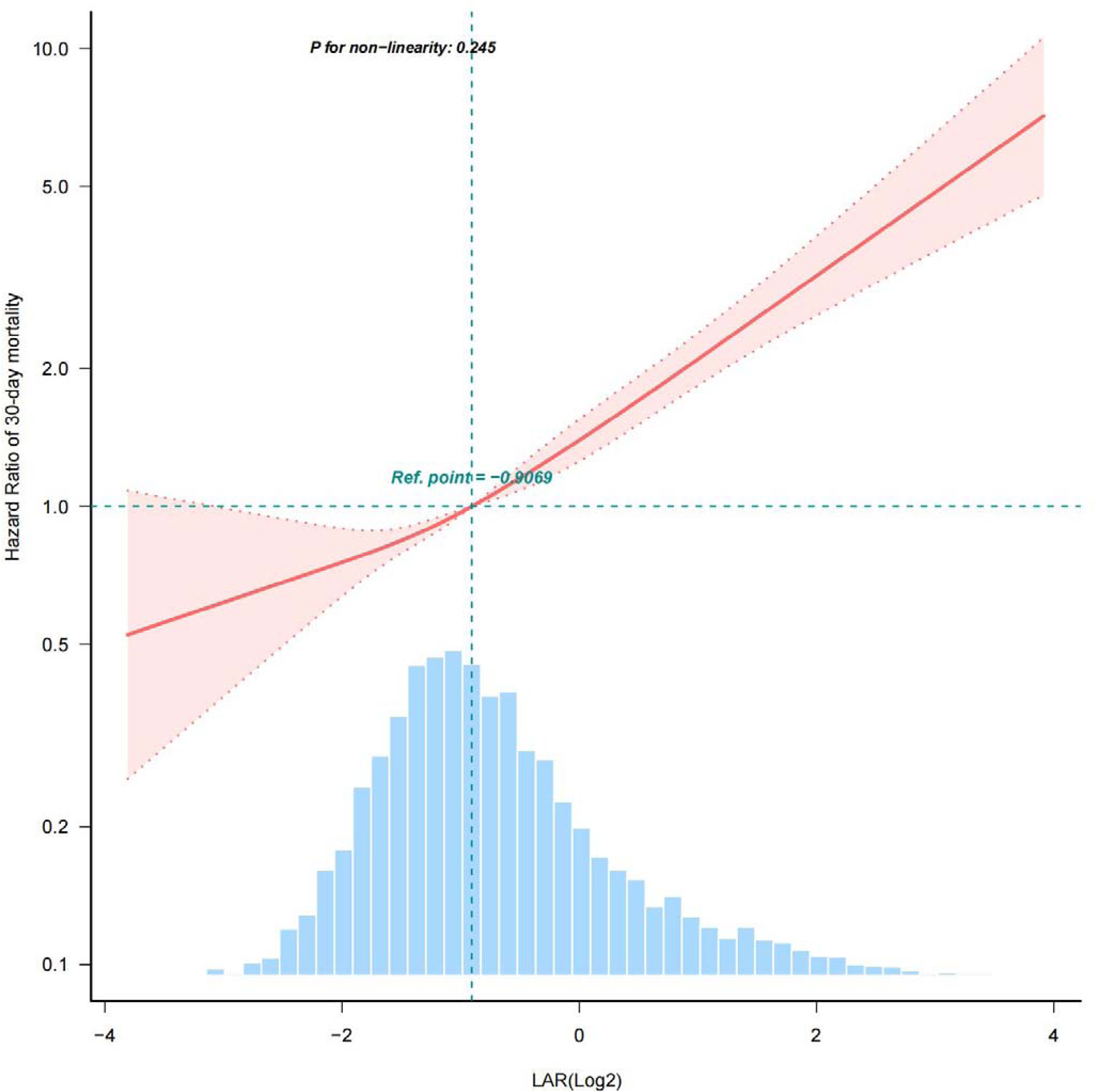
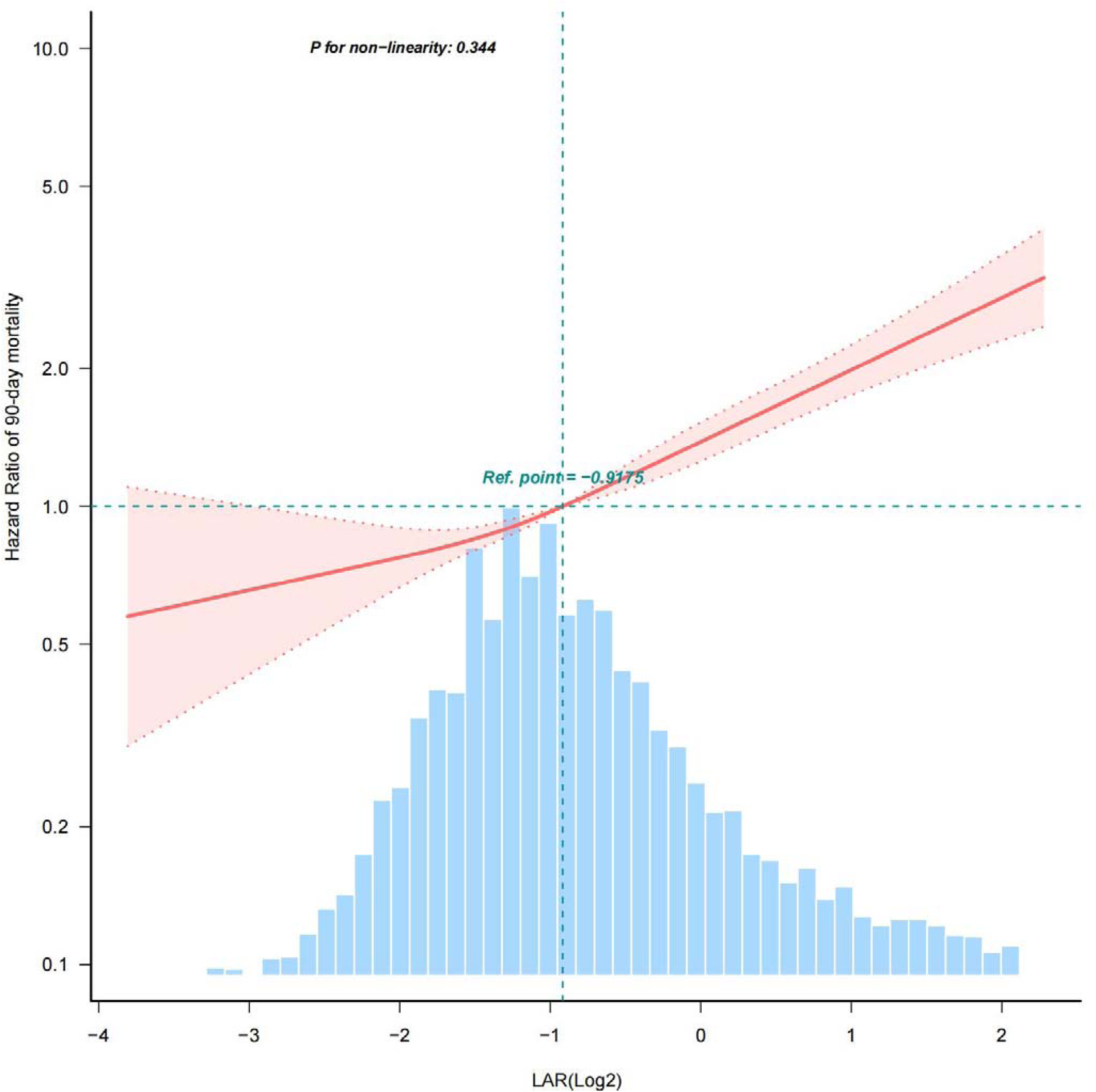
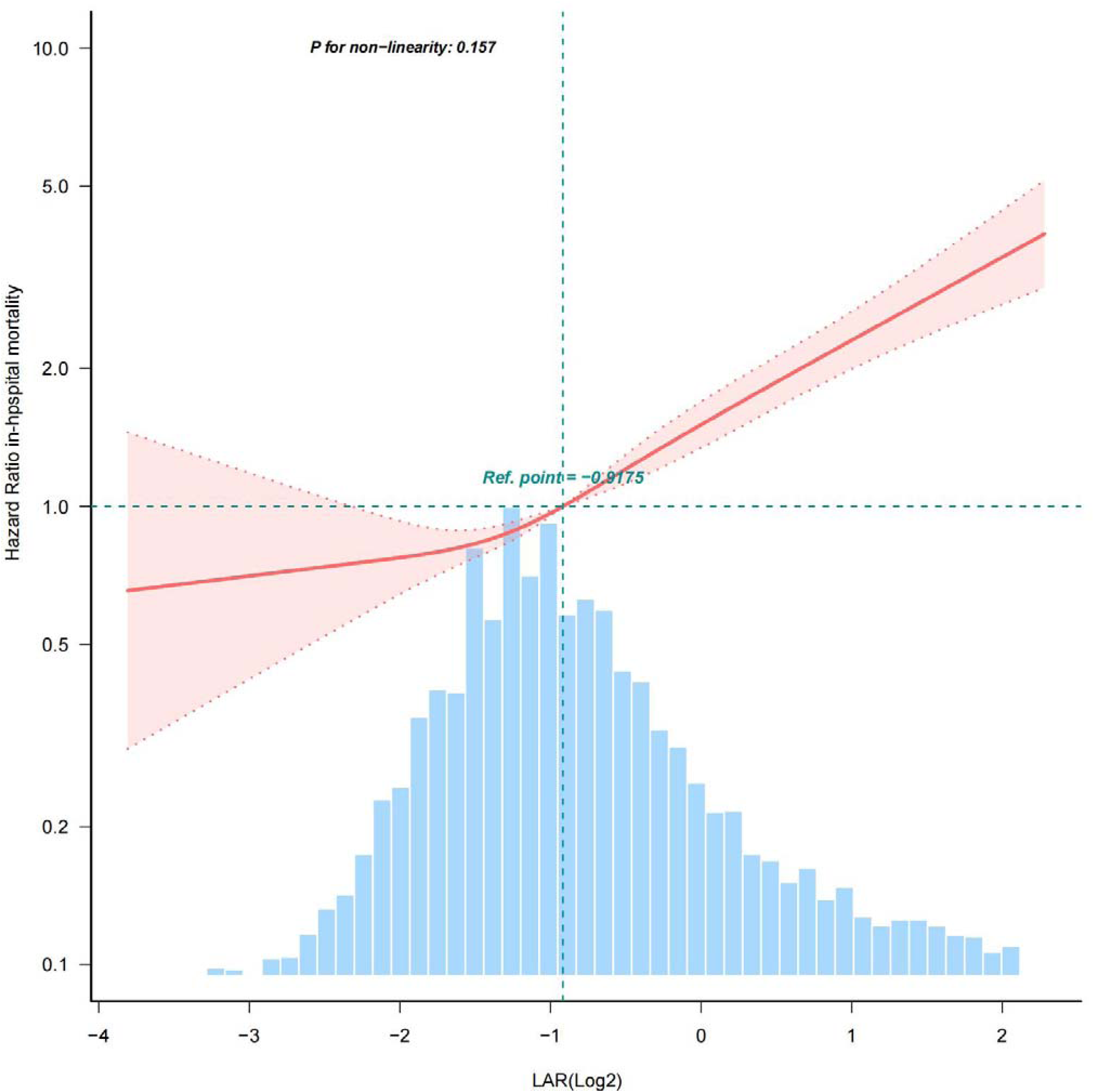

**Table 3.**
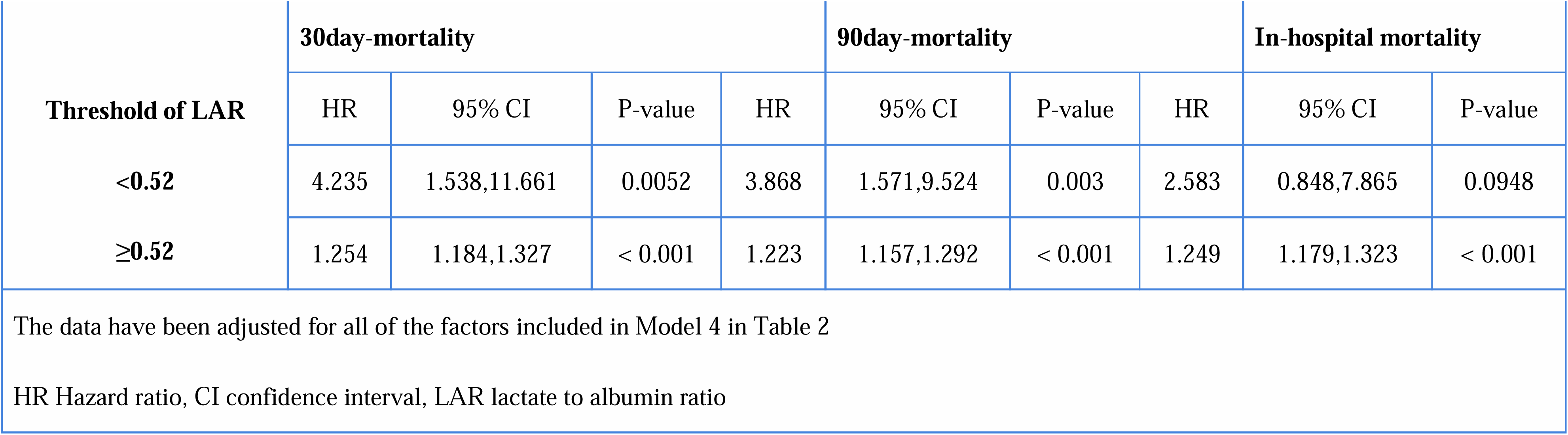
Threshold effect analysis of LAR on 30/90/in-hospital mortality in sepsis patients.

### Subgroup and sensitivity analysis

Subgroup analyses were used to determine the association between LAR and death in patients with sepsis **(Figure 3)**。We also performed stratified analyses according to age, sex, comorbidity, AKI stage, and sofa score. We found that the relationship between LAR and 30day,90day,inhospital mortality is fluctuating across partial subgroups. For incident 30day,90day,inhospital mortality, we observed significant differences in age, malignant cancerand AKI stage across three endings. In particular, mortality increased significantly with higher LAR when patients were younger than 65 years, or without malignancy, or AKI stage 1.

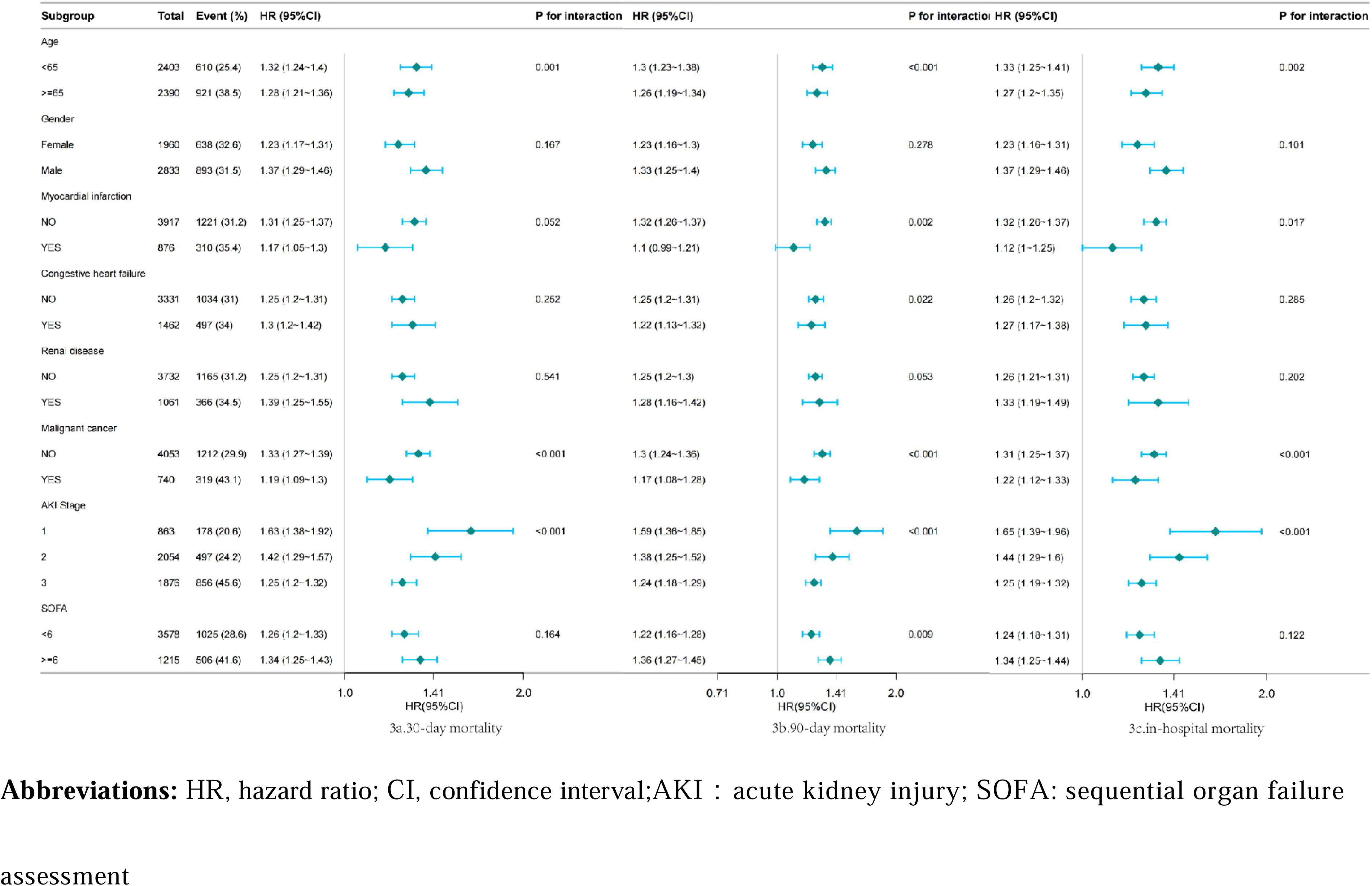

Second, when 90-day mortality was used as an endpoint, an interaction was observed in myocardial injury, congestive heart failure, as well as in the sofa subgroup, but it was not observed at 30 days and in-hospital death. In the myocardial injury subgroup, patients with myocardial injury had a low risk of 90 days relative to those with no myocardial injury (HR: 1.1,95%CI=0.99,1.21, HR: 1.32,95% CI = 1.26 to 1.37, respectively; p for interaction: 0.002). Interaction was also observed in the congestion heart failure subgroup (HR: 1.25,95% CI = 1.2-1.31, HR: 1.22,95%CI=1.13,1.32, respectively; p for interaction: 0.022). Similarly, patients with sofa <6 had a lower risk of 90 days compared to sofa 6 (HR: 1.22,95%CI=1.13,1.32, HR: 1.25,95% CI = 1.2 to 1.31, respectively; p for interaction: 0.009).

## DISCUSSION

In this retrospective cohort analysis of a large public dataset, we observed a significant association between LAR (Likelihood to Admit Ratio) and 30-day, 90-day, and in-hospital mortality in the S-AKI (Sepsis-Associated Acute Kidney Injury) population. COX multivariate regression analysis, subsequent to adjustments for confounding factors, consistently revealed a markedly heightened risk of mortality among S-AKI patients with LAR (log2).

Our study found a noteworthy association between LAR and S-AKI mortality. Recent studies suggest that LAR has predictive utility in mortality prediction in various diseases, including heart failure[15], sepsis[16] and acute respiratory failure[17]. Consistent with these insights, our study underscores the apparent association between elevated LAR and mortality. Our study found that the 30-day, 90-day and in-hospital mortality of patients with S-AKI was 31.9%, 38.2%, and 28.3%, which is consistent with supporting the high mortality associated with S-AKI[4]. Prior studies notably overlooked the comprehensive exploration of LAR’s impact on S-AKI. Our research broadens the scope of LAR’s relevance to a group of S-AKI patients, offering fresh perspectives within this particular medical context.

In our study, we evaluated the levels of lactate and albumin in the context of S-AKI. The findings revealed that elevated lactate levels, decreased albumin levels, and an increased LAR were linked to a higher risk of mortality in S-AKI patients.

This finding aligns with the emerging recognition of lactate and albumin as potential biomarkers for various pathological conditions. Lactate serves as an indicator of tissue hypoxia, while albumin is involved in the redox processes linked to different diseases. Previous research has shown a significant association between lactate levels and mortality in patients with suspected infection and sepsis[28]. In a study on COVID-19 mortality, the results similarly demonstrated a negative correlation, indicating higher mortality rates as albumin levels decreased. This underscores the potential importance of albumin as a prognostic marker in COVID-19 and potentially other disease conditions[18].

Using a multivariate regression analysis of LAR, we observed a higher risk of death in SAKI patients with increasing LAR. Studies have demonstrated that LAR has higher predictive accuracy for sepsis outcomes compared to individual assessments of lactate and albumin.A prospective study found that LAR was superior to lactate in predicting in-hospital mortality in sepsis[19]. Kamran Shadvar et al.[20] conducted a study involving 151 patients with septic shock. They found that LAR was superior to lactate levels and lactate clearance in predicting outcome in patients with septic shock. In a separate investigation by Cakir et al[21], including 1136 septic patients admitted to the ICU, LAR became a more reliable predictor of clinical outcome in patients with sepsis compared to individual assessments of lactate and albumin. These findings enhance our comprehension of the prognostic relevance of LAR in sepsis treatment but do not offer insight into S-AKI. Nevertheless, our study has established a significant link between LAR and 30-day, 90-day, and in-hospital mortality in the context of S-AKI. Additionally, the forest plot demonstrates a distinct interaction between age, AKI stage, and malignancy subgroups. These results offer valuable insights into the prognosis of S-AKI and the prognosis of specific population subgroups.

LAR is a potential independent risk factor for death in patients with S-AKI at 30,90 days as well as in-hospital days. Significant correlations were established by cox regression analysis and inflection point analysis and forest plots. The subgroup analysis in our study also indicated that significant interactions were seen across the three different observed endpoints when stratified by age, malignancy and AKI stage. However, because of our sample size limitations, especially the population reduction after stratification, the results may be biased and require further validation through large-scale prospective studies.

## LIMITATIONS

Our study has several limitations that should be considered. Firstly, as a single-center retrospective cohort study, the generalizability of our findings may be limited. Secondly, our focus on the relationship between the first LAR after admission and death in S-AKI patients may not fully reflect the relationship between dynamic LAR and mortality. Future studies should consider a more dynamic monitoring approach. Additionally, our dataset includes patients with comorbidities such as renal disease and malignancy, which may impact lactate and albumin levels. However, we conducted subgroup analyses for comorbidities and AKI stage, demonstrating the robustness of our results.

## CONCLUSION

In conclusion, our study indicates that LAR serves as an independent risk factor for 30-day, 90-day, and in-hospital mortality in S-AKI patients, with mortality increasing as LAR levels rise. However, further validation through large-scale multicenter prospective studies is necessary to establish its broad applicability and objectivity.

## Data Availability

The datasets generated and analyzed during the current study are available in the Medical Information Mart for Intensive Care IV (https://mimic.physionet. org/).

https://physionet.org/content/mimiciv/2.2/

## Acknowledgment

We are grateful to Dr. Jie Liu from the Department of Vascular and Endoascular Surgery of PLA General Hospital for his contribution to the statistical data.

## The contribution of the author

YW and HY contributed to the study conception, design, and revising the manuscript. YW participated in the data collection code extraction as well as in the analysis. Specifically, YW focused on the manuscript preparation, while HY revised the manuscript. In addition, HY is responsible for the project implementation and quality control. All authors read and approved the final draft.

## Declarations

### Ethics approval and consent to participate

The study was approved by Massachusetts Institute of Technology Affiliates. (ID: 52390976). The study conformed to the provisions of the Declaration of Helsinki (revised in 2013). The studies involving human participants were reviewed and approved by Institutional Review Board of the Massachusetts Institute of Technology and Beth Israel Deaconess Medical Center. The Medical Ethics Committee of Ningbo Medical Center Lihuili Hospital agreed to waive the ethics review because anonymous data was used in this study (ID: KY2023ML011).

### Consent for publication

Not applicable.

## REFERENCE

1. Fleischmann C, Scherag A, Adhikari NKJ, et al (2016) Assessment of Global Incidence and Mortality of Hospital-treated Sepsis. Current Estimates and Limitations. Am J Respir Crit Care Med 193:259–272. 10.1164/rccm.201504-0781OC

2. Singer M, Deutschman CS, Seymour CW, et al (2016) The Third International Consensus Definitions for Sepsis and Septic Shock (Sepsis-3). JAMA 315:801–810. 10.1001/jama.2016.0287

3. Rhee C, Dantes R, Epstein L, et al (2017) Incidence and Trends of Sepsis in US Hospitals Using Clinical vs Claims Data, 2009-2014. JAMA 318:1241–1249. 10.1001/jama.2017.13836

4. Liu J, Xie H, Ye Z, et al (2020) Rates, predictors, and mortality of sepsis-associated acute kidney injury: a systematic review and meta-analysis. BMC Nephrol 21:318. 10.1186/s12882-020-01974-8

5. A Prospective International Multicenter Study of AKI in the Intensive Care Unit - PubMed. https://pubmed.ncbi.nlm.nih.gov/26195505/. Accessed 10 May 2024

6. Septic acute kidney injury patients in emergency department: The risk factors and its correlation to serum lactate - PubMed. https://pubmed.ncbi.nlm.nih.gov/29776828/. Accessed 10 May 2024

7. Septic acute kidney injury in critically ill patients: clinical characteristics and outcomes - PubMed. https://pubmed.ncbi.nlm.nih.gov/17699448/. Accessed 10 May 2024

8. Severe hyperlactatemia, lactate clearance and mortality in unselected critically ill patients - PubMed. https://pubmed.ncbi.nlm.nih.gov/26556617/. Accessed 10 May 2024

9. Role of albumin in diseases associated with severe systemic inflammation: Pathophysiologic and clinical evidence in sepsis and in decompensated cirrhosis - PubMed. https://pubmed.ncbi.nlm.nih.gov/26831575/. Accessed 10 May 2024

10. Wang B, Chen G, Cao Y, et al (2015) Correlation of lactate/albumin ratio level to organ failure and mortality in severe sepsis and septic shock. J Crit Care 30:271–275. 10.1016/j.jcrc.2014.10.030

11. The Strengthening the Reporting of Observational Studies in Epidemiology (STROBE) statement: guidelines for reporting observational studies - PubMed. https://pubmed.ncbi.nlm.nih.gov/17938396/. Accessed 3 May 2024

12. MIMIC-IV, a freely accessible electronic health record dataset | Scientific Data. https://www.nature.com/articles/s41597-022-01899-x. Accessed 8 May 2024

13. Mervyn Singer, Singer M, Clifford S. Deutschman, et al (2016) The Third International Consensus Definitions for Sepsis and Septic Shock (Sepsis-3). JAMA 315:801–810. 10.1001/jama.2016.0287

14. Association Between Psoriasis and Nonalcoholic Fatty Liver Disease Among Outpatient US Adults - PubMed. https://pubmed.ncbi.nlm.nih.gov/35612851/. Accessed 13 Jan 2024

15. Guo W, Zhao L, Zhao H, et al (2021) The value of lactate/albumin ratio for predicting the clinical outcomes of critically ill patients with heart failure. Ann Transl Med 9:118. 10.21037/atm-20-4519

16. Lactate/albumin ratio is more effective than lactate or albumin alone in predicting clinical outcomes in intensive care patients with sepsis - PubMed. https://pubmed.ncbi.nlm.nih.gov/33745405/. Accessed 12 May 2024

17. Lu Y, Guo H, Chen X, Zhang Q (2021) Association between lactate/albumin ratio and all-cause mortality in patients with acute respiratory failure: A retrospective analysis. PLoS One 16:e0255744. 10.1371/journal.pone.0255744

18. Is Albumin Predictor of Mortality in COVID-19? - PubMed. https://pubmed.ncbi.nlm.nih.gov/32524832/. Accessed 12 May 2024

19. The prognostic value of the lactate/albumin ratio for predicting mortality in septic patients presenting to the emergency department: a prospective study - PubMed. https://pubmed.ncbi.nlm.nih.gov/34854770/. Accessed 12 May 2024

20. Shadvar K, Nader-Djalal N, Vahed N, et al (2022) Comparison of lactate/albumin ratio to lactate and lactate clearance for predicting outcomes in patients with septic shock admitted to intensive care unit: an observational study. Sci Rep 12:13047. 10.1038/s41598-022-14764-z

21. Cakir E, Turan IO (2021) Lactate/albumin ratio is more effective than lactate or albumin alone in predicting clinical outcomes in intensive care patients with sepsis. Scand J Clin Lab Invest 81:225–229. 10.1080/00365513.2021.1901306

